# Association between maternal lipid levels in third trimester and gestational diabetes mellitus among Vietnamese women

**DOI:** 10.1101/2025.05.22.25328151

**Authors:** Nguyen Viet Ha, Pham Manh Hung, Nguyen Tien Hoang, Pham Ba Nha

## Abstract

**Introduction:** Gestational diabetes mellitus (GDM) amplified the physiological alterations of maternal serum lipid levels in the third trimester, with higher triglycerides (TG) and lower high-density lipoprotein cholesterol (HDL-C) levels. No studies about the association between maternal lipid levels in the third trimester and the risk of GDM in Vietnamese population.

**Method:** a cross-sectional study on 1022 healthy females with singleton pregnancy delivered at Bach Mai hospital from May 2023 to June 2024. Measure fasting serum total cholesterol (TC), low-density lipoprotein cholesterol (LDL-C), high-density lipoprotein cholesterol (HDL-C) and triglycerides at 28 – 40 weeks of gestation by color enzymatic assays. Assess the association between maternal serum lipid levels with GDM by multivariate logistic regression.

**Results:** After adjusting for maternal age, pre-pregnancy body mass index (BMI), history of GDM, history of macrosomic neonates and family history of diabetes mellitus, every unit elevation of TG level was associated with increased risk of GDM (OR=1.25, 95% CI: 1.07 – 1.47); every unit elevation of LDL-C was associated with decreased risk of GDM (OR=0.61; 95%CI: 0.40 – 0.95). The association of LDL-C was significant in 33-36 weeks group (OR=0.35, 95% CI: 0.14 – 0.86). TG was significantly associated with GDM in 37 – 40 weeks group (OR=1.72; 95% CI: 1.32 – 2.25). To predict GDM, optimal cut-off points of LDL-C were ≤3.17 (for the third trimester) and ≤3.25 (for 33 – 36 weeks) while the threshold of TG were ≥3.30 (for the third trimester) and ≥3.31mmol/L (for 37 – 40 weeks).

**Conclusion:** Among Vietnamese population, LDL-C and TG were independent associated factors of GDM in the third trimester. Elevated level of TG was a risk factor, whereas low level of LDL-C was a protective factor of GDM.

## 1. Introduction

According to American Diabetes Association, gestational diabetes mellitus (GDM) is a type of diabetes diagnosed in the second or third trimester of pregnancy that was not clearly overt diabetes prior to gestation^1^. Its prevalence were 14% of pregnant women worldwide and 21,8% in Vietnam, based on International Association of Diabetes in Pregnancy Study Group’s Criteria and continue to rise due to increasing prevalence of obesity and sedentary lifestyle^2^. The risk of gestational diabetes mellitus increased in pregnant women with obesity, a history of macrosomic neonates, previous GDM, a family history of diabetes in a first-degree relative, or high-risk ethnicity^3^. If left untreated, the pregnant women with GDM may face short-term complications including gestational hypertension, cesarean section, preterm birth. Their newborns have higher rates of macrosomia, birth trauma, neonatal hyperbilirubinemia, hypoglycemia and respiratory distress syndrome^4^. Postnatally, GDM is associated with reoccurrence in subsequent pregnancies, cardiovascular diseases and type 2 diabetes in future^5^. Infants of GDM-complicated mothers face higher risk of metabolic syndrome, early-onset cardiovascular diseases, obesity, and hyperinsulinemia^6^.

The pathogenesis of GDM involves pancreatic insufficiency to compensate for elevated insulin resistance, secondary to hormonal changes accompanied with pregnancy progress. Insulin resistance in the second half of gestation results in the metabolic shift including reduced glucose consumption and increased lipolysis in order to reserve glucose for the fetus by placental transportation^7^. In response to hyperglycemia induced by insulin resistance, there is a rise of insulin secretion from pancreatic β-cells in normal pregnancy. However, any deficiencies in the insulin signaling process (pro-insulin synthesis, post-translational modifications, and gene alterations associated with insulin signaling) can result in β-cell dysfunction^8^. Chronic inflammation and the imbalance between oxidative stress and antioxidants also exacerbate both insulin resistance and β-cell dysfunction mechanisms in GDM^9^.

Previous studies showed the associations between GDM and dyslipidemia in pregnancy. GDM amplified the physiological alterations of maternal serum lipid levels in pregnancy which normally increase 30 to 50% for cholesterol and 200 to 400% for triglycerides by the end of gestation^10^. Maternal triglycerides (TG) and high-density lipoprotein cholesterol (HDL-C) level were two consistent lipid parameters associated with GDM in all trimesters, as evidenced by a systematic review and meta-analysis by Hu et al^11^. Pregnant women with GDM were more likely to have higher TG and lower HDL-C levels. In contrast, the associations of total cholesterol (TC) and low-density lipoprotein cholesterol (LDL-C) with GDM were changeable during pregnancy. GDM patients had significantly higher TC and LDL-C levels in the first trimester, but these relationships were inconsistent in the third trimester. Jin et al. also presented cut-off levels of TG (≥ 3,871mmol/L), HDL-C (≤ 1,712mmol/L) and LDL-C (≤ 2,415 mmol/L) for predicting GDM in late gestation^12^.

There have been no studies about maternal lipid levels in the third trimester and the association between them and the risk of GDM in Vietnamese population. Our study aims to investigate whether maternal lipid profiles, including TC, TG, HDL-C, LDL-C in third trimester are independently associated with the risk of GDM among Vietnamese pregnant women.

## II. Subjects and methods

### Study design

This cross-sectional study was conducted in Bach Mai Hospital, Hanoi, Vietnam. Bach Mai Hospital is a leading tertiary-level general hospital in northern Vietnam and its Department of Obstetrics and Gynecology is responsible for providing comprehensive care for women during pregnancy, childbirth, and the postpartum period. The department managed an average of 3,000 deliveries annually.

#### Participants

Participants included healthy pregnant women aged 18 – 45 years who attended prenatal care and intended to deliver at the department of Obstetrics and Gynecology of Bach Mai Hospital. These women were recruited between May 8^th^ 2023 and June 20^th^, 2024, using convenient sampling. Participants in the study, after being introduced to the research and invited to participate, will provide their consent by signing a consent form and writing “I agree to participate in the research.” No minors are involved in our study. Inclusion criteria were: 1) Gestational age 28 – 40 weeks; 2) Singleton pregnancy; 3) Natural conception. The participants were excluded if they had pre-existing chronic diseases, including hypertension, diabetes mellitus, thyroid, kidney, or cardiovascular diseases. Women with a history of tobacco or alcohol exposure during pregnancy, inherited metabolic diseases, or fetal anomalies were also excluded. We recruited 1022 pregnant women based on inclusion and exclusion criteria.

#### Data collection

At study entry, all recruited participants were requested to complete a questionnaire regarding maternal age, height, pre-pregnancy weight, residence, history of obstetric complications and family history of diabetes in first-degree relatives. Blood samples were collected to measure concentrations of TC, LDL-C, HDL-C, and TG. At delivery, we obtained data regarding gestational weight gain (GWG), HDP, delivery mode, gestational age, newborn sex, birthweight, 5-minute-postpartum Apgar score.

#### Biochemical analyses

Venous blood samples were taken after overnight fasting. Lipid profiles including TC, LDL-C, HDL-C and TG were measured using homogenous enzymatic colorimetric assays on an automatic biochemical analyzer (Beckman-Coulter AU5800, Beckman-Coulter Inc., California, USA) in the laboratory of Biochemistry Department of Bach Mai Hospital. The laboratory was accredited to ISO 15189: 2012 standards by the Bureau of Accreditation, Ministry of Science and Technology of Vietnam.

#### Definitions

Body mass index (BMI) was calculated by dividing weight in kilograms by the square of height in meters. Maternal pre-pregnancy BMI was categorized into underweight (<18.5 kg/m^2^), normal weight (18.5-24.9 kg/m^2^), overweight (25.0-29.9 kg/m^2^), and obese (≥30.0 kg/m^2^) groups according to World Health Organization BMI classification^13^.

GWG was stratified into appropriate, excessive and inadequate groups based on American Institute of Medicine^14^, in which appropriate GWG was defined as 12.5-18.0 kg in underweight women, 11.5-16.0 kg in normal weight women, 7.0-11.5 kg in overweight women and 5.0-9.0 kg in obese women. GWG greater than the thresholds were defined as excessive, while lower the thresholds was defined as inadequate GWG.

We used criteria from International Association of Diabetes and Pregnancy Study Groups to determine GDM with one-step oral glucose tolerance test performed at 24 – 28 weeks of gestation. GDM was diagnosed if any of the following venous plasma glucose thresholds are reached or surpassed: fasting 5.1 mmol/L, 1-hour post-load 10.0 mmol/L, and 2-hour post-load 8.5 mmol/L.

Newborn weight was classified as appropriate for gestational age, SGA and LGA based on standards from the International Fetal and Newborn Growth Consortium for the 21st Century (INTERGROWTH-21^st^). Neonates were defined as SGA if their birth weights fell under the 10^th^ percentile and LGA if their birth weights exceeded the 90^th^ percentile for gestational age and sex. AGA was defined as birth weights between the 10^th^ and 89^th^ percentile for gestational age and sex. Newborns were considered macrosomic if their birth weights were 4000 grams or more. Complicated pregnancy was defined as having at least one complication including HDP, GDM, preterm birth, SGA, LGA and macrosomia.

#### Statistical analysis

We presented normally and non-normally distributed continuous variable as mean±standard deviation (SD) and median (interquartile range, IQR), respectively. Categorical variables were presented as N (%). The difference of serum lipid levels between two non-normally distributed groups was tested using the Mann-Whitney U test, while Kruskal-Wallis test was used for the difference in three groups or more. Univariate and multivariate binary logistic regression was conducted to analyze the associations between serum TC, LDL-C, HDL-C and TG level and GDM in the third trimester. We considered advanced maternal age, pre-pregnancy overweight and obesity, history of GDM, history of macrosomic neonate and family history of DM in first-degree relatives as confounding variables. Then we used multivariate binary logistic regression to determine whether in each group of gestational age (28 – 32 weeks, 33 – 36 weeks and 37 – 40 weeks) the serum lipid levels were associated with the risk of GDM. Receiver Operating Characteristic (ROC) curve analysis was conducted to find the optimal cut-off points of serum lipid levels for predicting GDM in the third trimester and each group of gestational age. The cut-off point was considered as optimal if the corresponding Youden index (sensitivity + specificity – 1) reached maximum. Area under the curve (AUC) was calculated to describe the predicting power.. All analyses were performed with SPSS version 25.0 for Windows (SPSS Inc., Chicago, USA). P-value <0.05 were defined as statistically significant.

#### Ethical consideration

The study protocol was approved by Hanoi Medical University Institutional Ethical Review Board (IRB00002121) (Certificate of Approval No. 833/GCN-HÐÐÐNCYSH-ÐHYHN). All participants provided written informed consent after receiving a clear and comprehensive explanation of the study’s objectives, procedures, potential risks and benefits, and their right to withdraw at any time without penalty.

## Results

Maternal and neonatal characteristics are described in table 1. Mean ± SD of maternal age was 29.85 ± 4.90 with approximately 70% participants between 25 and 34 years old. About 8% of participants were overweight or obese while 14% of them were underweight. The percentage of women with history of macrosomic neonates and HDP were 2.4 and 1.7%, respectively. The distribution of gestational age at examination was relatively even across 3 groups with mean ± SD was 34.36 ± 2.77. Excessive GWG took up 18.4% of participants and the proportion of inadequate GWG was remarkable at 39.6%. The prevalence of HDP and GDM were 3.1 and 26.5%, respectively. Regarding neonates, the mean gestational age at delivery was 38.79 weeks with preterm percentage was 4.6%. Low birth weight and macrosomia accounted for 2.8 and 2.3%, respectively. Stratified for gestational age at delivery and newborn gender, 4.0% of them were SGA, 88.8% were AGA and 7.2% were LGA.

**Table 1.**
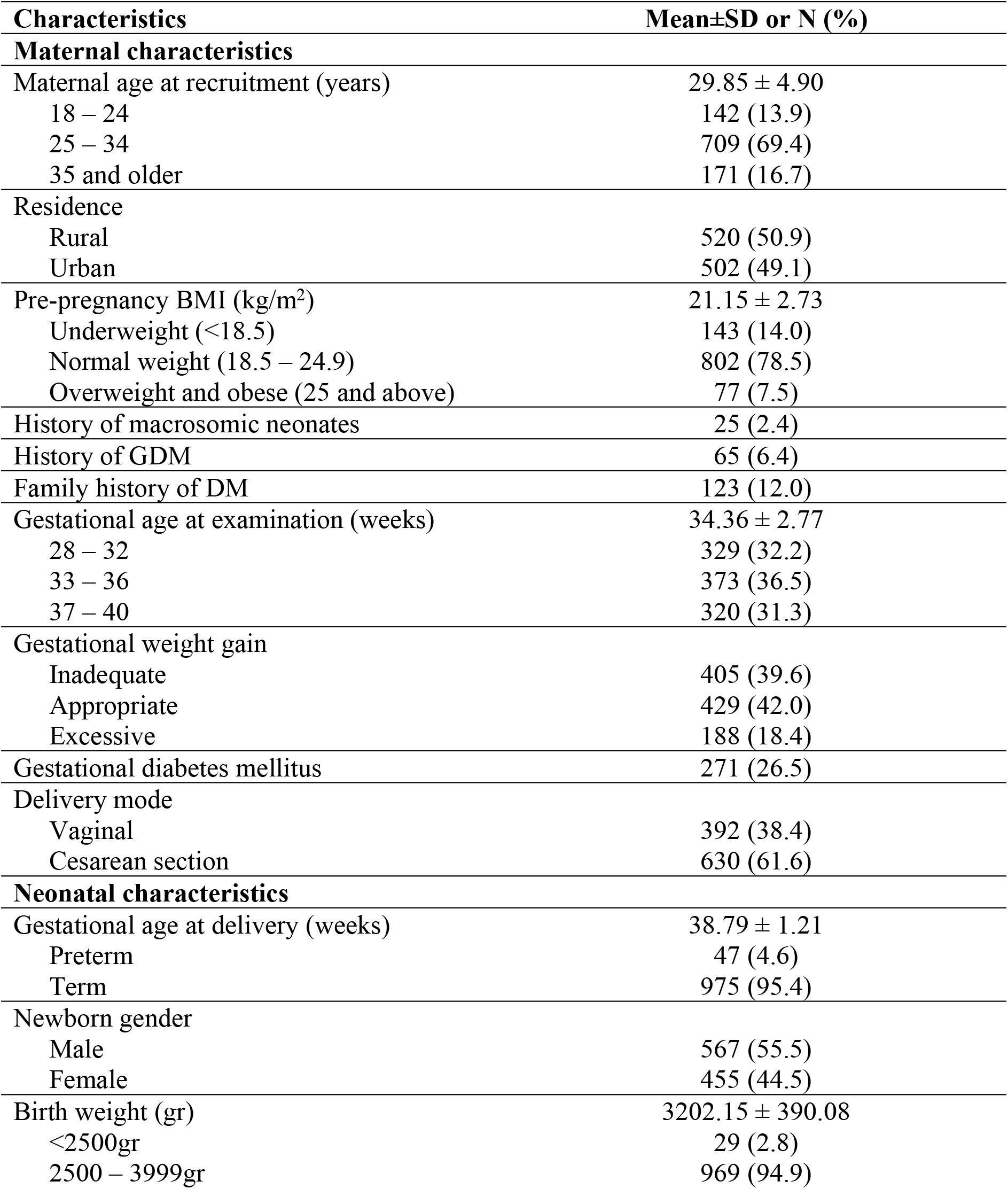
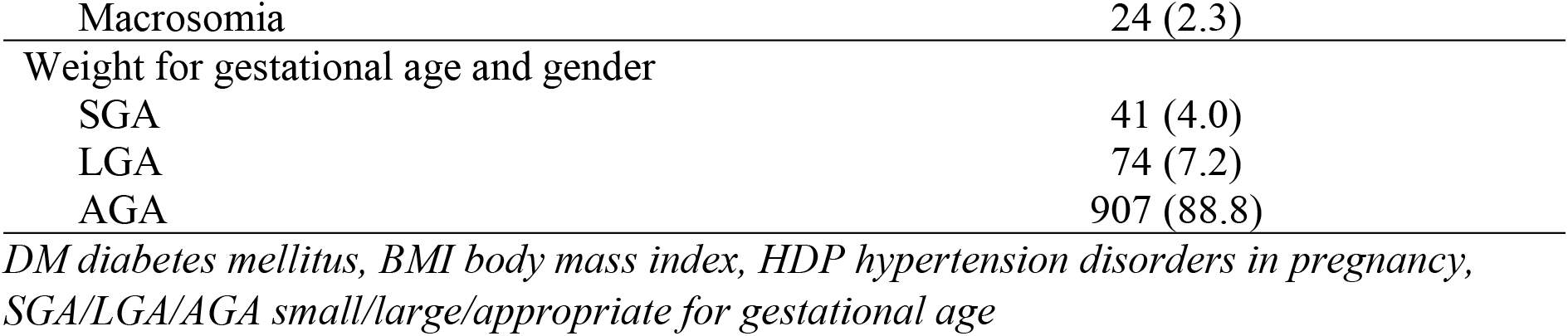
Maternal and neonatal characteristics.

Table 2 shows factors influencing maternal lipid concentrations in the third trimesters. Pre-pregnancy BMI was associated with the changes of all lipid levels, in which the higher degree of obesity the lower levels of TC, LDL-C, HDL-C and the higher level of TG. Regarding gestational age at examination, serum lipid levels increased progressively as the pregnancy advanced, with the exception of HDL-C. Women with previous macrosomic neonates had significantly higher level of TG than those without this history. Family history of DM in first-degree relatives was a factor related to significantly lower levels of TC and LDL-C in the third trimesters. GDM was associated with significant decrease in maternal TC, LDL-C and HDL-C as well as increase in TG concentrations.

**Table 2.** Factors associated with changes of maternal lipid levels in the third trimester.

| **Factors** | **TC** | **LDL-C** | **HDL-C** | **TG** |
| --- | --- | --- | --- | --- |
| **Maternal age (years)\*** |  |  |  |  |
| 18 – 24 | 6.56 (1.53) | 3.52 (1.16) | 1.79 (0.41) | 2.78 (1.14) |
| 25 -34 | 6.52 (1.50) | 3.33 (1.29) | 1.81 (0.41) | 2.89 (1.25) |
| 35 and older | 6.37 (1.52) | 3.28 (1.41) | 1.83 (0.40) | 3.00 (1.33) |
| p-value | 0.39 | 0.08 | 0.66 | 0.12 |
| **Gestational age at examination\*** |  |  |  |  |
| 28 – 32 weeks | 6.34 (1.60) | 3.30 (1.46) | 1.83 (0.47) | 2.77 (1.18) |
| 33 – 36 weeks | 6.35 (1.50) | 3.30 (1.13) | 1.81 (0.40) | 2.87 (1.15) |
| 37 – 40 weeks | 6.68 (1.45) | 3.44 (1.29) | 1.82 (0.37) | 3.06 (1.49) |
| p-value | **<0.01** | **0.02** | 0.54 | **<0.01** |
| **Pre-pregnancy BMI (kg/m<sup>2</sup>)\*** |  |  |  |  |
| <18,5 | 6.63 (1.62) | 3.65 (1.40) | 1.85 (0.40) | 2.65 (1.13) |
| 18,5 – 24,9 | 6.49 (1.51) | 3.30 (1.29) | 1.81 (0.41) | 2.91 (1.29) |
| ≥ 25 | 6.27 (1.61) | 3.19 (1.40) | 1.72 (0.43) | 3.12 (1.22) |
| p-value | **0.01** | **<0.01** | **0.02** | **<0.01** |
| **History of GDM\*\*** |  |  |  |  |
| Yes | 6.35 (1.74) | 3.20 (1.64) | 1.77 (0.45) | 2.88 (1.44) |
| No | 6.49 (1.51) | 3.35 (1.30) | 1.82 (0.41) | 1.90 (1.25) |
| p-value | 0.58 | 0.50 | 0.44 | 0.39 |
| **History of macrosomic neonates\*\*** |  |  |  |  |
| Yes | 6.15 (1.25) | 3.08 (1.20) | 1.78 (0.55) | 3.46 (2.00) |
| No | 6.49 (1.52) | 3.36 (1.30) | 1.81 (0.41) | 2.88 (1.25) |
| p-value | 0.06 | 0.07 | 0.50 | **0.01** |
| **Family history of DM\*\*** |  |  |  |  |
| Yes | 6.24 (1.31) | 3.10 (1.11) | 1.77 (0.48) | 2.93 (1.48) |
| No | 6.52 (1.51) | 3.38 (1.32) | 1.82 (0.40) | 2.88 (1.24) |
| p-value | **0.01** | **0.01** | 0.10 | 0.35 |
| **GDM\*\*** |  |  |  |  |
| Yes | 6.32 (1.66) | 3.07 (1.37) | 1.74 (0.45) | 3.26 (1.50) |
| No | 6.55 (1.47) | 3.43 (1.28) | 1.83 (0.38) | 2.82 (1.13) |
| p-value | **0.03** | **0.01** | **0.01** | **0.01** |

\*Compare differences by Kruskal-Wallis test
\*\*Compare differences by Mann-Whitney test.

The result of univariate and multivariate analysis for association between GDM and maternal lipid panel in the third trimester was demonstrated in Table 3. Univariate logistic regression revealed inverse association of TC, LDL-C and HDL-C as well as positive association of TG concentrations with GDM in the third trimester. After adjusting for maternal age, pre-pregnancy BMI, history of GDM, history of macrosomic neonates and family history of DM, only LDL-C and TG were significantly associated factors. Particularly, every mmol/L elevation of LDL-C was inversely related to GDM risk, with 39% decrease, while every mmol/L elevation of TG was positively related, with 1,25-fold increase.

**Table 3.** Binary logistic analysis of factors associated with GDM.

| Factors | Crude OR (95% CI) | p-value | Adjusted OR (95% CI) | p-value |
| --- | --- | --- | --- | --- |
| **Maternal age** |  |  |  |  |
| ≥ 35 | 2.17 (1.54 – 3.07) | **0.01** | 1.75 (1.21 – 2.54) | **0.01** |
| <35 | 1 |  | 1 |  |
| **Pre-pregnancy BMI** |  |  |  |  |
| ≥25 | 2.10 (1.31 – 3.38) | **0.01** | 1.75 (1.05 – 2.91) | **0.03** |
| <25 | 1 |  | 1 |  |
| **History of GDM** |  |  |  |  |
| Yes | 3.81 (2.29 – 6.36) | **0.01** | 3.08 (1.79 – 5.30) | **0.01** |
| No | 1 |  | 1 |  |
| **History of macrosomic neonates** |  |  |  |  |
| Yes | 3.67 (1.64 – 8.18) | **0.01** | 2.46 (1.03 – 5.87) | **0.04** |
| No | 1 |  | 1 |  |
| **Family history of DM** |  |  |  |  |
| Yes | 2.46 (1.67 – 3.62) | **0.01** | 1.80 (1.19 – 2.73) | **0.01** |
| No | 1 |  | 1 |  |
| **TC** | 0.84 (0.75 – 0.95) | **0.01** | 1.32 (0.87 – 2.02) | 0.19 |
| **LDL-C** | 0.75 (0.65 – 0.82) | **0.01** | 0.61(0.40 – 0.95) | **0.03** |
| **HDL-C** | 0.46 (0.29 – 0.72) | **0.01** | 0.57 (0.29 – 1.14) | 0.11 |
| **TG** | 1.38 (1.23 – 1.56) | **0.01** | 1.25 (1.07 – 1.47) | **0.01** |

We then explore whether the association between maternal serum lipid levels in the third trimester and GDM varies with gestational age (Table 4). No lipid concentrations at 28-32 weeks of gestation were found to be correlated with the risk of GDM. For gestation age 33 – 36 weeks, only LDL-C level had inverse association with GDM risk. The association between TG levels and GDM was observed exclusively in the 37-40 weeks of gestation.

**Table 4.** Association between lipid panel and GDM based on gestational age.

| Lipids | 28 – 32 weeks |  | 33 – 36 weeks |  | 37 – 40 weeks |  |
| --- | --- | --- | --- | --- | --- | --- |
|  | aOR (95% CI) | p-value | aOR (95% CI) | p-value | aOR (95% CI) | p-value |
| TC | 1.13<br>(0.52-2.47) | 0.75 | 2.13<br>(0.87-5.19) | 0.10 | 1.48<br>(0.74-2.95) | 0.27 |
| LDL-C | 0.83<br>(0.37-1.85) | 0.65 | 0.35<br>(0.14-0.86) | **0.02** | 0.49<br>(0.24-1.01) | 0.06 |
| HDL-C | 0.58<br>(0.19-1.78) | 0.34 | 0.37<br>(0.09-1.46) | 0.16 | 0.58<br>(0.16-2.10) | 0.40 |
| TG | 1.19<br>(0.86-1.64) | 0.29 | 1.03<br>(0.74-1.45) | 0.84 | 1.72<br>(1.32-2.25) | **0.01** |

*aOR: adjusted odd ratio. Confounding variables were advanced maternal age, pre-pregnancy overweight and obesity, history of GDM, history of macrosomic neonate and family history of DM in first-degree relatives.*

Table 5 presents the optimal cut-off points of maternal lipids levels for predicting GDM in the third trimester and groups of gestational age. In the third trimester, cut-off points of LDL-C and TG were ≤ 3.17 and ≥ 3.30 mmol/L, respectively. TG had greater predicting power compared to LDL-C based on larger AUC. Stratification by gestational age enhanced the predicting power of both lipids. In 33 – 36 weeks group, LDL-C ≤ 3.25 mmol/L had better AUC, sensitivity, and specificity than in third trimester. In 37 – 40 weeks group, similar improvements were observed for TG with the threshold of 3.31 mmol/L, compared to in the third trimester.

**Table 5.**
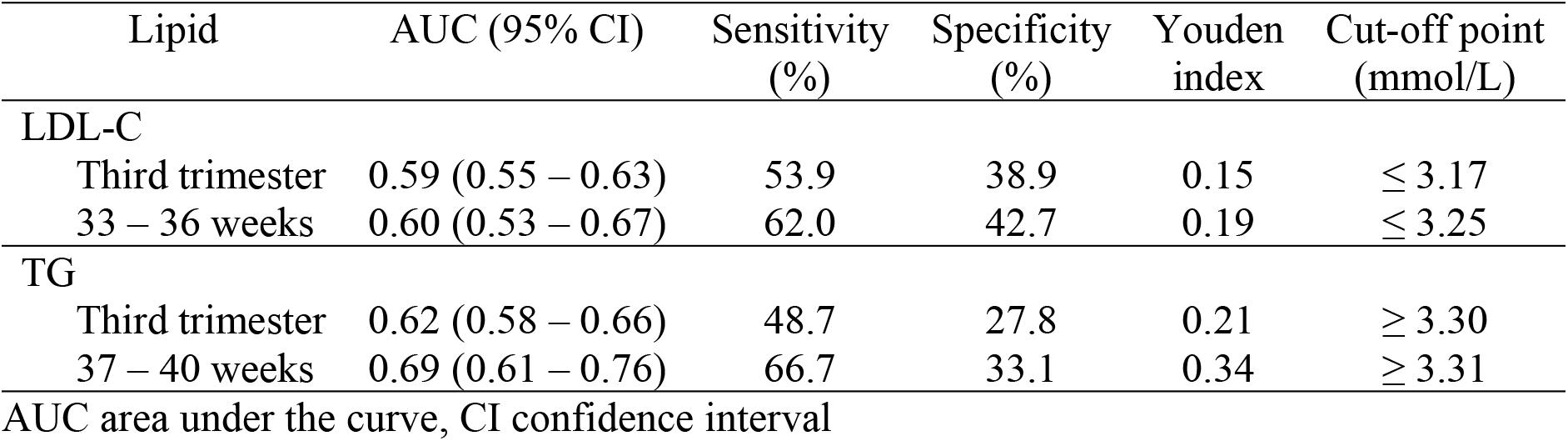
Cut-off points of maternal third trimester lipids for predicting GDM.

| Lipid | AUC (95% CI) | Sensitivity (%) | Specificity (%) | Youden index | Cut-off point (mmol/L) |
| --- | --- | --- | --- | --- | --- |
| LDL-C |  |  |  |  |  |
| Third trimester | 0.59 (0.55 – 0.63) | 53.9 | 38.9 | 0.15 | $\leq 3.17$ |
| 33 – 36 weeks | 0.60 (0.53 – 0.67) | 62.0 | 42.7 | 0.19 | $\leq 3.25$ |
| TG |  |  |  |  |  |
| Third trimester | 0.62 (0.58 – 0.66) | 48.7 | 27.8 | 0.21 | $\geq 3.30$ |
| 37 – 40 weeks | 0.69 (0.61 – 0.76) | 66.7 | 33.1 | 0.34 | $\geq 3.31$ |

AUC area under the curve, CI confidence interval

## IV. Discussion

Our prospective cross-sectional study comprehensively investigated the association between the risk of GDM and third-trimester serum TC, LDL-C, HDL-C, and triglycerides among Vietnamese pregnant women. We also explored these associations in each month of the third trimester and presented the cut-off points of maternal lipids for GDM prediction.

Our research findings indicated high TG and low LDL-C levels during the third trimester were associated with an elevated risk of GDM. This correlation between maternal TG and GDM was consistent with the result of a large meta-analysis^11^. Several studies on Chinese population also presented the association between GDM and low LDL-C in the third trimester^12,15^ while the result from Hu’s meta-analysis only showed higher LDL-C in GDM patients, especially during the first trimester^11^. As lipid levels gradually elevate during the third trimester, we assessed these associations in each month of the third trimester and explored that the association of LDL-C and of TG only presented in group of 33 – 36 weeks and 37 – 40 weeks, respectively. Consequently, predicting power of LDL-C and TG for GDM was improved when applied to the corresponding group. These are novel findings of this study, suggesting the complexity of maternal lipid changes in the pathology of GDM.

There are two phases of maternal lipid changes during pregnancy including anabolic and catabolic phase. The former one is observed in the first half of the pregnancy characterized by maternal fat accumulation due to increased maternal appetite and food intake^16^ as well as minimal change in insulin sensitivity^17^. The latter phase is presented with elevated serum TC, LDL-C and particularly TG levels until delivery. HDL-C concentration peaked in the 7th month and remained unchanged by the time of parturition. These changes result from elevated insulin resistance in which placental hormones^18^ (estrogen, progesterone, cortisol, prolactin and human placental lactogen) and adipokines^19^ (leptin, tumor necrosis factor-α, adiponectin and resistin) contribute to. The gradual increase in maternal lipid levels during the third trimester, except for HDL-C, found in our study is consistent with previous reasearch^20,21^. Thus, gestational age is a key factor to evaluate maternal lipid concentrations in pregnancy. Besides, pre-pregnancy BMI, diet and ethnicity also significantly influence maternal lipid levels^22^.

The association between GDM and maternal changes in lipid levels is probably bi-directional. Lipid changes in early pregnancy, characterized by increased TC, LDL-C and TG along with decreased HDL-C, predispose the pregnant women to GDM development^11^. These changes indicate maternal insulin resistance in which there are increased lipolysis to release free fatty acids leading to increase triglycerides production in liver as well as augmented HDL breakdown and impaired HDL synthesis^23^. On the other hand, alterations of maternal lipid levels in the third trimester are secondary to GDM. Insulin deficiency in GDM is linked to elevated TG level, as insulin activates lipoprotein lipase to enhance TG clearance. Also, decreased LDL-C level were founded to be associated to GDM in few studies^12,15^. The implementation of a low-fat diet in GDM patients, known to reduce TC and LDL-C levels^24^, may contribute to this phenomenon. Another possible mechanism is elevated activity of cholesteryl ester transfer protein triggered by high TG level. Consequently, the exchange of cholesterol ester within LDL with TG in TG-rich lipoproteins is augmented^25^, which results in reduced levels of LDL-C.

Given the impact of gestation age on maternal serum lipid levels during pregnancy, we analyzed the association between lipid concentrations and GDM by three gestational age groups. Using this approach, we found associations between GDM and LDL-C and TG that only presented in the specific gestational age group. Also, implementing gestational age-specific thresholds for LDL-C and TG in GDM prediction enhanced the predicting power of these biomarkers. Our study presented a lower cut-off point of TG and a higher one of LDL-C than in Jin’s study^12^. Several factors may account for these differences. Beside ethnicity, pregnant women in Jin’s study had a lower incidence of GDM (7.8%) compared to our study. Moreover, the distribution of gestational age in Jin’s cohort was not mentioned. Last but not least, both studies did not collect data regarding participants’ diet, maybe due to difficulties measuring and assessing food intake by interview.

There are some strengths and limitations in our study. We conducted the first research on Vietnamese population regarding lipid levels in third trimester. The large sample size, with relatively equal distribution across gestational age groups in the third trimester, is a significant strength of this study. To our knowledge, few studies in third trimester provided the distribution of gestational age. We did not only analyze the association of lipid levels and GDM in the third trimester but also in each month of it. Resultantly, we presented detailed cut-off points for LDL—C and TG for predicting GDM. Besides, all blood samples were measured within 2 hours after sampling on one system. This method helped avoid the possible impact of measurement and the freeze-thaw process.

However, several limitations should be noted. First, a single measurement of serum lipid level may lead to misclassification of maternal status. Second, the maternal pre-pregnancy weight, history of disease, and other basic data were self-reported, and recall bias is difficult to avoid. Third, some lifestyle factors such as behavior and physical activity, which are related to lipid levels, were not collected. Finally, this is a single-center study with limited population diversity. A multi-center study with multiple populations across Vietnam would provide more representative results.

## V. Conclusion

In conclusion, we explored the associations between GDM and maternal serum LDL-C and TG levels in the third trimester after adjusting for other confounders. Among three groups of gestational age, there were significant associations of LDL-C and TG in 33 – 36 weeks and 37 – 40 weeks, respectively. We also presented the cut-off points to predict GDM in the third trimester (LDL-C ≤ 3.17 and TG ≥ 3.30 mmol/L). For gestational age 33 – 36 weeks, cut-off point of LDL-C was ≤ 3.25mmol/L while the threshold for TG was ≥ 3.31mmol/L for 37 – 40 weeks.

## Data Availability

Data cannot be shared publicly because of research information security regulations of Bach Mai Hospital and Hanoi Medical University. Data are available from the Institutional Data Access / Ethics Committee (contact via) for researchers who meet the criteria for access to confidential data.

## CRediT Authorship Contribution Statement

NVH conceptualization, methodology, investigation, formal analysis, writing – original draft. PMH methodology, resources, visualization, writing – review and editing. NTH supervision, validation and writing – review and editing. PBN methodology, validation, supervision, writing – review and editing.

## References

1. American Diabetes Association Professional Practice C. 2. Diagnosis and Classification of Diabetes: Standards of Care in Diabetes—2024. Diabetes Care. 2023;47(Supplement_1):S20–S42.

2. Wang H, Li N, Chivese T, et al. IDF Diabetes Atlas: Estimation of Global and Regional Gestational Diabetes Mellitus Prevalence for 2021 by International Association of Diabetes in Pregnancy Study Group’s Criteria. Diabetes Research and Clinical Practice. 2022;183:109050.

3. National Institute for Health and Care Excellence: Guidelines. In: Diabetes in pregnancy: management from preconception to the postnatal period. London: National Institute for Health and Care Excellence (NICE); 2020.

4. 2. Classification and Diagnosis of Diabetes: Standards of Medical Care in Diabetes-2018. Diabetes Care. 2018;41(Suppl 1):S13–s27.

5. Farahvar S, Walfisch A, Sheiner E. Gestational diabetes risk factors and long-term consequences for both mother and offspring: a literature review. Expert Review of Endocrinology & Metabolism. 2019;14(1):63–63.

6. Sweeting A, Wong J, Murphy HR, Ross GP. A Clinical Update on Gestational Diabetes Mellitus. Endocr Rev. 2022;43(5):763–763.

7. Choudhury AA, Devi Rajeswari V. Gestational diabetes mellitus - A metabolic and reproductive disorder. Biomedicine & Pharmacotherapy. 2021;143:112183.

8. Plows JF, Stanley JL, Baker PN, Reynolds CM, Vickers MH. The Pathophysiology of Gestational Diabetes Mellitus. International Journal of Molecular Sciences. 2018;19(11). doi:10.3390/ijms19113342.

9. Shamsad A, Kushwah AS, Singh R, Banerjee M. Pharmaco-epi-genetic and pathophysiology of gestational diabetes mellitus (GDM): An overview. Health Sciences Review. 2023;7:100086.

10. Basaran A. Pregnancy-induced Hyperlipoproteinemia: Review of the Literature. Reproductive Sciences. 2009;16(5):431–431.

11. Hu J, Gillies CL, Lin S, et al. Association of maternal lipid profile and gestational diabetes mellitus: A systematic review and meta-analysis of 292 studies and 97,880 women. EClinicalMedicine. 2021;34:100830.

12. Jin W-Y, Lin S-L, Hou R-L, et al. Associations between maternal lipid profile and pregnancy complications and perinatal outcomes: a population-based study from China. BMC Pregnancy and Childbirth. 2016;16(1):60.

13. Obesity WHOCo, World Health O. Obesity : preventing and managing the global epidemic : report of a WHO consultation. In. Geneva: World Health Organization; 2000.

14. American College of Obstetrics and Gynecology. ACOG Committee opinion no. 548: weight gain during pregnancy. Obstet Gynecol. 2013;121(1):210–210.

15. Zhang Y, Lan X, Cai C, et al. Associations between Maternal Lipid Profiles and Pregnancy Complications: A Prospective Population-Based Study. Am J Perinatol. 2021;38(8):834–834.

16. Murphy SP, Abrams BF. Changes in energy intakes during pregnancy and lactation in a national sample of US women. Am J Public Health. 1993;83(8):1161–1161.

17. Skajaa GØ, Fuglsang J, Kampmann U, Ovesen PG. Parity increases insulin requirements in pregnant women with type 1 diabetes. The Journal of Clinical Endocrinology & Metabolism. 2018;103(6):2302–2302.

18. Ramírez SI, Suniega EA, Laughrey MI. Endocrinology During Pregnancy. Primary Care: Clinics in Office Practice. 2024;51(3):535–535.

19. Mallardo M, Ferraro S, Daniele A, Nigro E. GDM-complicated pregnancies: focus on adipokines. Mol Biol Rep. 2021;48(12):8171–8171.

20. Lockitch G, Gamer PR. Clinical Biochemistry of Pregnancy. Critical Reviews in Clinical Laboratory Sciences. 1997;34(1):67–67.

21. Wiznitzer A, Mayer A, Novack V, et al. Association of lipid levels during gestation with preeclampsia and gestational diabetes mellitus: a population-based study. Am J Obstet Gynecol. 2009;201(5):482.e481–488.

22. Mulder JWCM, Kusters DM, Roeters van Lennep JE, Hutten BA. Lipid metabolism during pregnancy: consequences for mother and child. Current Opinion in Lipidology. 2024;35(3).

23. Baneu P, Văcărescu C, Drăgan SR, et al. The Triglyceride/HDL Ratio as a Surrogate Biomarker for Insulin Resistance. Biomedicines. 2024;12(7).

24. Khoury J, Henriksen T, Christophersen B, Tonstad S. Effect of a cholesterol-lowering diet on maternal, cord, and neonatal lipids, and pregnancy outcome: A randomized clinical trial. American Journal of Obstetrics and Gynecology. 2005;193(4):1292–1292.

25. Barter PJ, Brewer HB, Chapman MJ, Hennekens CH, Rader DJ, Tall AR. Cholesteryl Ester Transfer Protein. Arteriosclerosis, Thrombosis, and Vascular Biology. 2003;23(2):160–160.

